# The Lebanese Cohort for COVID-19; A Challenge for the ABO Blood Group System

**DOI:** 10.1101/2020.08.02.20166785

**Authors:** Athar Khalil, Rita Feghali, Mahmoud Hassoun

**Affiliations:** Department of Biochemistry and Molecular Genetics, American University of Beirut, Beirut, Lebanon; Department of Laboratory Medicine, Rafik Hariri University Hospital, Beirut, Lebanon; Department of Pulmonary and Intensive Care Unit, Rafik Hariri University Hospital, Beirut, Lebanon

**Keywords:** SARS-CoV-2, COVID-19 cohort, ABO blood group system, Infection predisposition

## Abstract

A sudden outbreak of pneumonia caused by the Severe Acute Respiratory Syndrome Coronavirus 2 (SARS-CoV-2) has rapidly spread all over the world facilitating the declaration of the resultant disease as a pandemic in March,2020. In Lebanon, the fast action of announcing a state of emergency with strict measures was among the factors that helped in achieving a successful containment of the disease in the country. Predisposing factors for acquiring COVID-19 and for developing a severe form of this disease were postulated to be related to epidemiological and clinical characteristics as well as the genomics signature of a given population or its environment. Biological markers such as the ABO blood group system was amongst those factors that were proposed to be linked to the variability in the disease course and/or the prevalence of this infection among different groups. We therefore conducted the first retrospective case-control study in the Middle-East and North Africa that tackles the association between the blood group types and the susceptibility as well as the severity of SARS-CoV2 infection. Opposing to the current acknowledged hypothesis, our results have challenged the association significance of this system with COVID-19. Herein, we highlighted the importance of studying larger cohorts using more rigorous approaches to diminish the potential confounding effect of some underlying comorbidities and genetic variants that are known to be associated with the ABO blood group system.

## 1. Introduction

In December 2019, a sudden outbreak of pneumonia has emerged in Wu Han City, China that then spread all over the world^1^. This coronavirus disease 2019 (COVID-19) as named by the World Health Organization(WHO), is caused by the Betacoronavirus SARS-CoV-2 that targets primarily the human’s lower respiratory tract. COVID-19 is mainly characterized by dry cough, dyspnea, fever, and bilateral lung infiltrates upon imaging. The primary cause of death among the infected patients remains to be the acquired severe respiratory failure ^2^. The fast spread of this contagious disease reaching around 200 countries in a short period of time has facilitated the declaration of COVID-19 outbreak as a pandemic on March 11^th^ 2020^3^. Till this date, the global number of confirmed positive cases exceeded the 10 million and caused more than 500,000 deaths^4^. In Lebanon, the official authorities had forced strict measures shortly after detecting the first case of COVID-19. This fast action of declaring a state of emergency in the country was among the factors that helped in the successful containment of the disease. So far, the Ministry of Public Health has confirmed 1885 cases of COVID-19 and 36 associated deaths ^5,6^. As such, Lebanon has avoided the best-case projection scenario, according to China modeling, that anticipated tens of thousands cases to be diagnosed in our region. Comparing to other countries with small population like Norway which reported till this date around 8,936 confirmed cases with 251 deaths from a total population of 5.3 million, Lebanon did a good job in managing the spread of this disease ^4,7^.

Yet, it should be noted that the predisposition for acquiring COVID-19 and for developing severe form of this disease is also known to be affected by some epidemiological and clinical characteristics of the population. The main studied risk factors are age, sex, and some chronic conditions such as diabetes and cardiovascular diseases^8,9^. On the contrary, the establishment of biological markers for predicting the variability in the disease course or the prevalence of infection among different groups is not yet elucidated^10^. From that perspective and due to the highly tackled association between blood groups and the susceptibility/severity of various diseases such as SARS-CoV-1, P. falciparum, H. pylori, Norwalk virus, hepatitis B virus, and N. gonorrhoeae, investigations on its association with COVID-19 cases was evaluated^11^. Zhao et.al were the first to report an association between ABO blood group and the susceptibility to SARS-CoV-2^12^. Afterwards, several studies has emerged to pinpoint again on the increased risk for SARS-CoV-2 infection and the higher mortality rate among blood group A individuals as compared to that of blood group O^10–13^. The explanation of this linkage was attributed to several hypothesis such as the presence of an extra sugar N-acetyl galactosamine on the surface of blood group A cells that can possibly propose a more pathogen-host contact or the presence of anti-A antibodies IgGs in the serum of O blood type carriers that can inhibit the virus–cell adhesion process (Figure-1)^13,14^. Recently, a newly published article in The New England Journal of Medicine presented a genetic piece of evidence that established a potential role of the ABO blood-group system as a risk factor for acquiring COVID-19. In their article, Ellinghaus et. al have identified the rs657152 variant that coincided on the ABO blood group locus (9q34.2) to be associated with COVID-19–induced respiratory failure^15^.

Although all the above mentioned articles provided a consistent association between the ABO blood system and COVID-19 disease, yet, all of these studies emphasized on the importance of investigating this association in larger clinical datasets among different populations. For this reason, we decided to evaluate the association between different blood types and the susceptibility as well as severity of SARS-CoV2 infection in the Lebanese Cohort.

**Figure 1:**
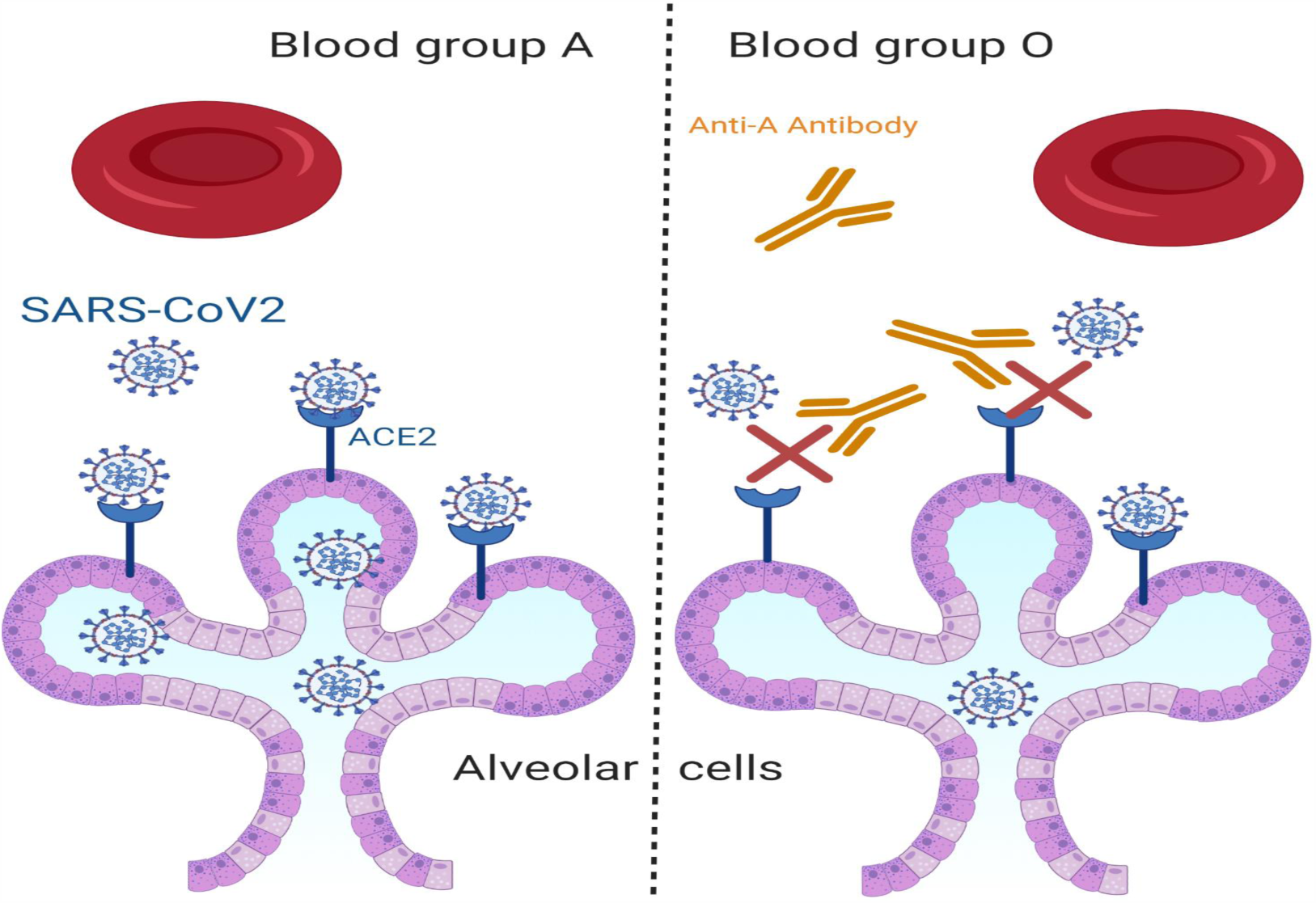
One of the proposed hypothesis for ABO blood group association with COVID-19. Increased risk for SARS-CoV-2 infection among blood group A individuals as compared to that of blood group O due to the presence of anti-A antibodies in the serum of O blood type carriers that can inhibit the virus– cell adhesion process.

## 2. Methods

### a. Data Source

Medical records for COVID-19 patients and their clinical characteristics were obtained from the electronic medical database at Rafik Hariri University Hospital (RHUH). The retrieved variables for COVID-19 patients were age, sex, blood group and severity of their disease. For control population, only blood group types ware retrieved.

### b. Study design

This is a retrospective case-control study with 146 COVID-19 patients and 6497 participants in the control group. The case group consisted of individuals with known blood types who were diagnosed with COVID-19 and hospitalized in RHUH between February, 2020 and June, 2020. The diagnosis of COVID-19 was confirmed by a positive real-time reverse transcriptase polymerase-chain-reaction test for SARS-CoV-2 on nasal and pharyngeal swab specimens from patients. For the control group, blood groups data for all individuals who were hospitalized in RHUH between 01/01/2018 till 24/06/2020 were included. For the control group, all blood samples were typed by forward test tube method while for the case group the typing was done by both forward and reverse testing.

Severity classification criteria was adopted from the interim guidelines of the World Health Organization and the National Health Commission of China. Based on the patients’ symptoms, laboratory results and imaging findings at admission, the disease severity of COVID-19 patients was categorized into four types: mild, moderate, severe, and critical^16,17^. Given the small sample size of our cohort and the low number of severe and critical cases, we combined these two categories together. Briefly,1) Mild cases patients were those diagnosed with upper respiratory symptoms but with no imaging abnormalities 2) Moderate patients were diagnosed based on the presence of at least two of the following : dyspnea, cough, and body temperature > 38°C, with imaging abnormalities, 3) Severe cases were characterized by respiratory distress with respiratory rate ≥ 30 Breaths Per Minute (BPM), O2 Saturation ≤ 93%, and PaO2/FiO2 ≤ 300 mmHg while those categorized as 4) critical cases are those who had acute respiratory failure necessitating non-invasive or/and invasive ventilation with the presence of multi organ failure such as renal failure, liver failure and heart failure, shock and imaging with more than 50 % progression within 24-48 hours^16^.

This study was approved by the Ethical Committee of RHUH. Due to the retrospective nature of the study and because no identifying information relating to participants was included, written informed consent was waived. All experimental protocols were conducted according to the Strengthening of the Reporting of Observational Studies in Epidemiology guidelines.

### c. Statistical analysis

Statistical computations were performed using IBM SPSS statistics 24. Data were analyzed by t-test. χ2 test and Fisher’s exact test when applicable. We compared each blood group against all others using a 1×2 contingency table to determine effect sizes for each blood group in our COVID-19 cohort. For studying the significance of adjusted risk factors, grouping was done as following (mild versus moderate and severe/critical, severe/ critical versus mild and moderate) this allowed us to study the association of each blood group with the mild or severe/critical form of the disease compared to the rest of the cases. Binary logistic regression analysis adjusted for age and gender was done for each blood group versus the rest of blood groups. Results were reported as odds ratio with 95% confidence interval. P-values of less than or equal to 0.05 were considered statistically significant.

## 3. Results

### a. Patients’

clinical characteristics and the ABO blood group distribution among patients and control group

A total of 146 patients diagnosed with COVID-19 were enrolled in this study. The mean age was 41.9 years old (interquartile range (IQR), 28–57 years, p < 0.001) and those who are between 22 and 40 years old constituted the largest portion of our cohort (45.2%)(Figure-2). The frequency of male gender (n=97,66.4%) was significantly higher than that of the female (n=49,33.6%) (Table-1). Frequencies of blood groups showed that the highest percentage belong to blood group A (40.4%) followed by blood group O (35.6%) and then blood group B (17.1%) and AB (6.8%) (Table-1). Mild cases scored the highest percentage in this cohort were 72 patients (58.6%) belonged to this groups while those with severe or critical symptoms presented only 13.7% of the cases (Table-1).

**Figure 2:**
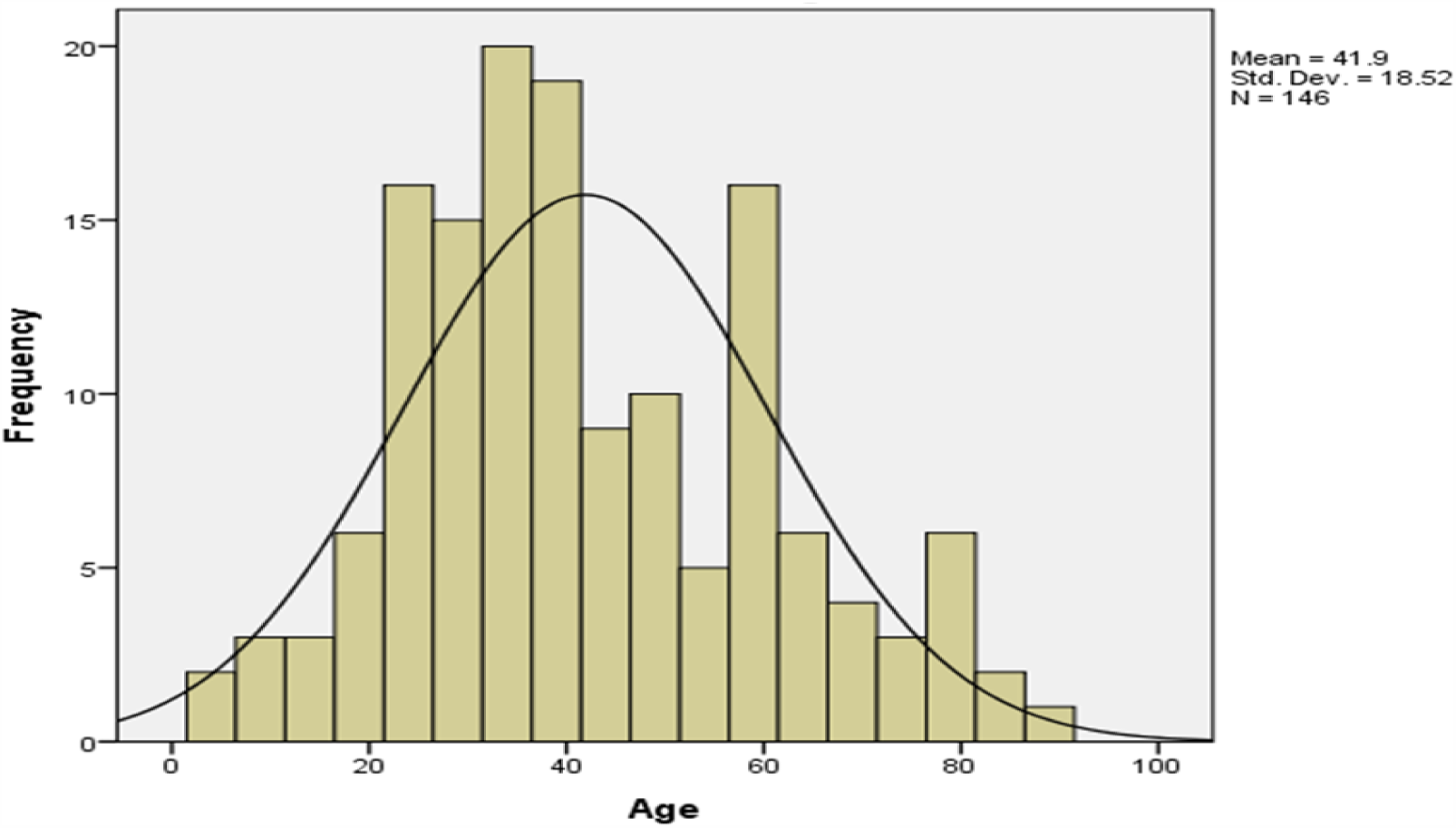
Age distribution pattern among our COVD-19 cohort. The mean age for the involved 146 patients in this study was 41.9 (t-test=27.336, p value < 0.001).

**Table 1:** Clinical characteristics of COVID-19 patients including sex, ABO blood groups and severity outcome of SARS-CoV2 infection.

| Variables | | Numbers | Percentages | $\chi^2$ | P-Value |
| --- | --- | --- | --- | --- | --- |
| **Sex** | Male | 97 | 66.4% | 15.781 | <.001 |
|  | Female | 49 | 33.6% |  |  |
| **Severity of COVID-19** | Mild | 72 | 49.3% | 28.658 | <.001 |
|  | Moderate | 54 | 37% |  |  |
|  | Severe/<br>Critical | 20 | 13.7% |  |  |
| **ABO blood group** | A | 59 | 40.4% | 43.315 | <.001 |
|  | AB | 10 | 6.8% |  |  |
|  | B | 25 | 17.1% |  |  |
|  | O | 52 | 35.6% |  |  |

Our control group constituted from a total of 6479 cases admitted to RHUH between 1/1/2018 and 1/6/2020. Blood group O was the highest detected blood group with 2573 cases (39.7%) while AB blood group scored the lowest number with 352 cases (5.4%). Although the percentage of patients with blood group A was higher among COVID-19 cases as compared to that of the control group (40.4% vs. 35.8%, P = 0.25) while that of blood group O was lower (35.6% vs. 39.7%, P = 0.32) yet, these differences did not reach statistical significance (Table-2).

**Table 2:** The ABO blood group distribution among COVID-19 patients and normal controls.

|  | COVID-19 patients | Control group |  |  |
| --- | --- | --- | --- | --- |
| **Subjects** | 146 (100%) | 6479 (100%) | $\chi^2$ | P-Value |
| **Blood group, n(%)** |  |  |  |  |
| **A** | 59 (40.4%) | 2322 (35.8%) | 1.2964 | .25487 |
| **B** | 25 (17.1%) | 1232(19%) | .3325 | .564215 |
| **O** | 52 (35.6%) | 2573(39.7%) | 1.0016 | .316932 |
| **AB** | 10(6.9%) | 352 (5.4%) | .5545 | .456478 |

### b. The effect of age, sex and blood group on dictating the severity of COVID-19

After stratifying our data based on the severity outcome of the disease, only the age factor showed a significant association with the three categories of severity (p<0.001). Mild cases were mostly patients below 40 years old, moderate cases were those between 40-59 years (46.3%) while severe and critical cases belonged mainly to those above 60 years old (55%). On the other hand, the association of severity with blood groups and gender didn’t reach a statistical significance. This is reasonable since in all the outcome categories, blood group A and male gender were the predominant (Table-3). Taken into consideration that age as an independent variable had a significant association with our outcome and since previous studies have revealed a potential effect of gender on the severity of the disease, we went further in our analysis by adjusting for age and gender. Blood groups and the severity outcomes were divided into two sets in each round of analysis to allow for a precise and specific association studies. As such, the risk factor for each blood group was studied separately for the mild and severe form of the disease while adjusting for the age and gender variables. Only individuals with blood group AB had higher odds for the mild form of the disease (OR=4.613, <0.108) but with borderline significance(Table-4).

**Table 3:**
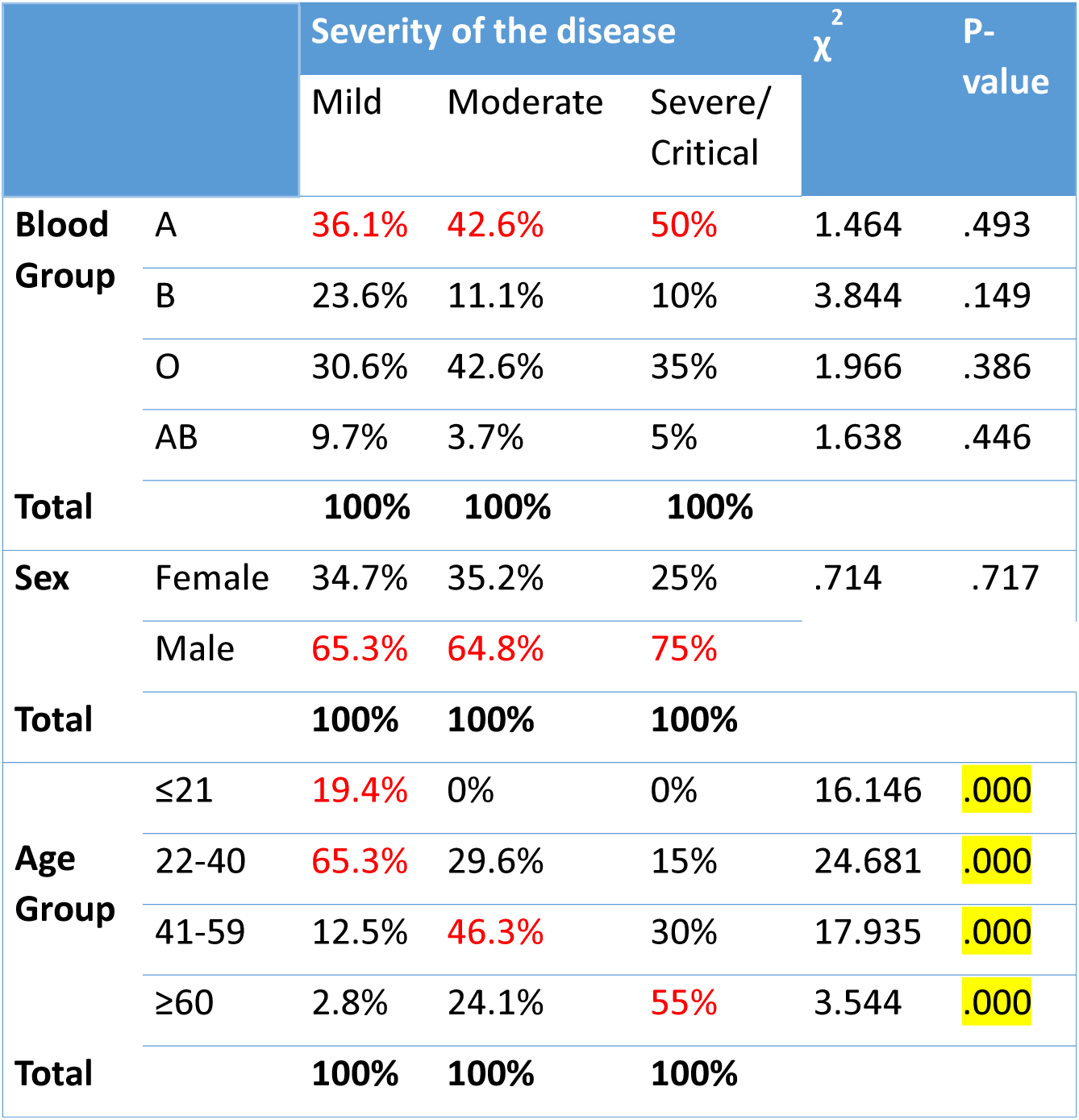
The unadjusted association between the stratified outcome of COVID-19 and various variables including age, sex and ABO blood groups.

| | | Severity of the disease | | | $\chi^2$ | P-value |
| --- | --- | --- | --- | --- | --- | --- |
|  |  | Mild | Moderate | Severe/<br>Critical |  |  |
| **Blood Group** | A | 36.1% | 42.6% | 50% | 1.464 | .493 |
|  | B | 23.6% | 11.1% | 10% | 3.844 | .149 |
|  | O | 30.6% | 42.6% | 35% | 1.966 | .386 |
|  | AB | 9.7% | 3.7% | 5% | 1.638 | .446 |
| **Total** |  | **100%** | **100%** | **100%** |  |  |
| **Sex** | Female | 34.7% | 35.2% | 25% | .714 | .717 |
|  | Male | 65.3% | 64.8% | 75% |  |  |
| **Total** |  | **100%** | **100%** | **100%** |  |  |
| **Age Group** | ≤21 | 19.4% | 0% | 0% | 16.146 | .000 |
|  | 22-40 | 65.3% | 29.6% | 15% | 24.681 | .000 |
|  | 41-59 | 12.5% | 46.3% | 30% | 17.935 | .000 |
|  | ≥60 | 2.8% | 24.1% | 55% | 3.544 | .000 |
| **Total** |  | **100%** | **100%** | **100%** |  |  |

**Table 4:** Logistic regression analysis adjusted by age and sex for assessing the effect of each blood group on COVID-19 outcome. OR= Odds Ratio, CI= Confidence interval, P= P value

| Outcome<br>ABO groups | Mild . VS. Not Mild | Severe .VS. Not Severe |
| --- | --- | --- |
| **A—Non A** | 26 (44.1%) | 10 (16.9%) |
|  | OR=1.029<br>CI: .414-2.559<br>P=.951 | OR= .752<br>CI: .238-2.378<br>P =.628 |
| **B—Non B** | 17 (68%) | 2 (8%) |
|  | OR= 2.847<br>CI: .905-9.959<br>P= .73 | OR= .879<br>CI: .170-4.538<br>P=.877 |
| **O—Non O** | 22 (42.3%) | 7 (13.5 %) |
|  | OR= .340<br>CI: .135-.855<br>P=.22 | OR= 1.536<br>CI.487-4.849<br>P=.464 |
| **AB—Non AB** | 7 (70%) | 1 (10%) |
|  | OR= 4.613<br>CI: .717-29.684<br>P= .108 | OR= .784<br>CI: .083-7.393<br>P= .832 |

## 4. Discussion

This retrospective study was done at the Rafik Hariri University Hospital (RHUH)– formerly known as Beirut Governmental University Hospital which is the largest Lebanese public hospital. Currently, RHUH is the leading center for COVID-19 testing and treatment in the country. Our aim was to analyze the distribution of the ABO blood groups among COVID-19 Lebanese citizens while taking into consideration that of the control group (n=6497). For our knowledge, this is the first study in the Middle East region that tackles the association between the ABO blood groups and the susceptibility to COVID-19 infection while accounting for the population blood group distribution pattern. Globally, we are the first to question the significance of ABO blood system as an independent risk factor for the stratified outcomes of the disease.

Similar to results from previous studies, blood group A was the most prevalent in our COVID-19 cohort (40.4%, P <0.001)^18^. This association lost it significance when the distribution of blood groups among the control group was taken into consideration. In our study we have reassured the previously established pattern of the ABO blood groups distribution in our population where blood group O is presented as the predominant blood type ^19^. Since the distribution of blood groups among COVID-19 patients showed a comparable scattering pattern with that of the general population, the association significance between blood groups and the infection susceptibility was not distinguished^20^. In this regard, it should be well-noted that some of the previously conveyed data did not account for the general population distribution for blood groups while those who took into consideration this calculation involved low representing numbers that might not be sufficient for a significant reflection to the entire population^10,18,21^. Moreover, discrepancies between the conveyed results prevent the firm agreement on a specific blood group to be a risk factor for acquiring SARS-CoV-2. Although initially blood group A was proposed as a risk factor, subsequent studies from Iran, Saudi and Shenzhen, proposed that AB individuals are rather at higher risk to acquire this disease as compared to other blood groups^18,22^. This inconsistent association could be explained by the different epidemiological patterns among various ethnicities which pinpoint on the importance of including larger cohorts for more precise associations and conclusions.

So far, multiple risk factors for developing a severe form of SARS-CoV2 infection are established. Our data have re-assured the significance of age in enhancing the severity of the infection among patients. Age factor is confirmed to be associated with poor outcomes characterized by severe symptoms with non-improvement and higher mortality rates^17,23^. This was explained by the fact that the strong host innate immune responses among older individuals can cause an insufficient control of viral replication and a prolonged pro-inflammatory responses which can in turn lead to a marked decline in cell-mediated and humoral immune function^17^. For gender distribution, the higher infection incidence among men in our cohort was similar to the globally accepted pattern. But when stratified by disease severity, male gender was not considered as a significant risk factor for developing a severe form of the disease which is consistent with the previously published results^24^.

Since ABO blood groups and/or other comorbidities such as cardiovascular diseases were speculated to be prognostic markers for COVID-19 and not risk factors predisposing SARS-CoV-2 infection, we studied the association of this system with COVID-19 outcome.^25^ Herein, we were unable to report a significant association between any of the blood groups and the severity outcome among COVID-19 patients even when other influencing factor were taken into consideration. Data from several other cohorts have already revealed a non-significant association between the ABO system and the severity, ICU admission, intubation and death among patients ^10,11,22^. Additionally, the recent GWAS results revealed a non-significant difference in the blood-group distribution between patients receiving only supplemental oxygen and those receiving mechanical ventilation of any kind^15^. Thus, our results could be added to the list that question the significance of presenting blood group system as a risk factor for dictating COVID-19 outcome. One suggested explanation is that once SARS-CoV2 infection is fully established, it can replicate in the individual’s epithelial cells by exhibiting the individual’s antigen, and thus rendering the individual’s ABO antibodies ineffective against the newly produced viruses.

The flood of fast-paced research and reporting milieu concerning this pandemic, especially when dealing with preprint servers, is causing confusions and is triggering controversy information. Major pitfalls in COVID-19 research concerning the involvement of the blood group system were identified. The analysis of the data in most of the published studies have neglected the effect of other risk factors and comorbidities that are known to worsen SARS-coV2 infection and that can be distributed differently among blood groups. For example, blood group A is considered a significant risk factor for coronary heart disease (CHD), venous deep thromboembolism (VTE), fever, cough, dyspnea, sore throat, chest pain/distress and fatigue which are all implicated in COVID-19 prognosis ^21,26,27^. A recent study have established a significant association between some COVID-19 commodities/variables and blood group A but these cofounders were not adjusted when studying the independent effect of blood groups on the severity outcome^10^. Similarly, allele A lost its significance as a risk factor for susceptibility and mortality in COVID-19 when polymorphisms of angiotensin-converting enzyme 1 (ACE1), complement component 3 (C3) were added to the multivariate regression model^28^. While we acknowledge that the main limitation of our study is the sample size, we are the first to evaluate the association between different blood groups with the stratified outcome of COVID-19 while taking into consideration the gender and age factors. In conclusion, larger cohorts with more rigorous approaches are recommended to diminish the potential confounding effect of some underlying comorbidities and genetic variants, which are known to be associated with the ABO blood group and can be over-presented in COVID-19 cohorts^26^.

## Data Availability

NA

## Author contributions

AK, RF and MH contributed to the conception or design of the work. AK contributed to the acquisition, analysis, or interpretation of data for the work. AK drafted the manuscript. AK, RF, and MH critically revised the manuscript. All gave final approval and agree to be accountable for all aspects of work ensuring integrity and accuracy.

## Acknowledgements

The authors would like to thank Dr. Firas Abiad the general manager at Rafik Hariri University Hospital were this study took place. The schematic illustration was created using Biorender.com.

## Conflict of Interest

The authors declare that the research was conducted in the absence of any commercial or financial relationships that could be construed as a potential conflict of interest.

## Notes

### Competing Interest Statement

The authors have declared no competing interest.

